# Association between resting heart rate and all-cause mortality in patients with intracerebral hemorrhage in the intensive care unit: a retrospective study based on the MIMIC-IV database

**DOI:** 10.1101/2024.12.31.24319835

**Authors:** Xinwei Yao, Junyu Yang, Tao Zhang, Ying Pang, Chunyu Zhang, Dan Huang, Tongjie Ji, Siyi Xu, Min Liu, Chunglong Zhong

## Abstract

**Background:** Intracerebral hemorrhage (ICH) is a leading cause of disability and mortality, accounting for 20-30% of acute cerebrovascular events. Resting heart rate (RHR) is an important vital sign linked to prognosis, particularly in cardiovascular diseases. This study investigates the association between RHR and all-cause mortality (ACM) in intensive care unit (ICU) patients with ICH.

**Methods:** Data were extracted from the MIMIC-IV database. Patients were divided into quartiles (Q1–Q4) based on RHR. ACM at 30 days, 90 days, and one year was assessed. Kaplan-Meier curves, multivariate Cox regression, and restricted cubic splines were used to analyze the relationship between RHR and ACM, with receiver operating characteristic curves evaluating the predictive value of RHR >90.3 beats per minute (bpm).

**Results:** A total of 1,918 patients were included. Survival curves showed significantly higher mortality in higher RHR quartiles (Q3 and Q4) compared to Q1 and Q2. Multivariate Cox regression confirmed that RHR above the second quartile (RHR=78.5 bpm) was linked to higher mortality. Restricted cubic splines revealed a nonlinear relationship between elevated RHR and increased mortality.

**Conclusions:** A resting heart rate below 90.3 bpm may serve as an independent protective factor against all-cause mortality in patients with intracerebral hemorrhage in the intensive care unit. In contrast, an elevated resting heart rate exceeding 90.3 bpm may be independently associated with an increased risk of all-cause mortality in this population.

## Introduction

According to the Global Burden of Disease 2021 (GBD 2021) Stroke Risk Factor Collaborators, stroke is the second most common cause of mortality and the third most common cause of long-term disability globally in 2021[1]. Meanwhile intracerebral hemorrhage (ICH), a subtype of hemorrhagic stroke in a specific pathological classification that is characterized by bleeding into the brain parenchyma with the formation of a hematoma[2], [3], accounts for approximately 20%-30% of all strokes and contributes to about 40% of deaths from stroke events[1], [4]. The mortality rate of patients diagnosed with ICH may be as high as 50%, and the rates of complications from ICH-related dysfunction are reported to be 32.8%-42.4% at 6 months and 16.7%-24.6% at 12 months post-ICH [5][6]. The high mortality and morbidity associated with ICH, along with the increasing stroke burden from 1990 to 2021, particularly in certain regions such as East Asia, Central Asia, and sub-Saharan Africa, have made ICH a significant threat to global health[1].

In recent years, an increasing number of biomarkers and novel mechanisms including immunological responses and ferroptosis in ICH, have been identified and highlighted[3], [7], [8]. This has contributed to the growing consensus that ICH may be more than just a localized event, but rather a systemic disease with broader pathological implications. The central nervous system (CNS) has extensive physiologic effects on the cardiovascular system[9], [10]. When brain injury occurs, pathological conditions of the cardiovascular system can be detected, including elevated cardiac marker levels, arrhythmias, abnormal electrocardiogram results, myocardial necrosis, and autonomic dysfunction[11–13]. Dysfunction of the cardiovascular system such as auricular fibrillation-related acute stroke, similarly affects the CNS[11]. Neurocardiology has been highlighted as a subspecialty focused on advancing our understanding of the complex interactions between the brain and the heart through innovative research. It also aims to foster integrated care and promote collaboration between neurologists and cardiologists, with the ultimate goal of improving clinical outcomes[14–16].

Heart rate, as one of the most fundamental and easily obtainable vital signs in the ICU, and its neurocardiac influence has been explored. It was suggested in a study of Rass V [17] that a greater variation in heart rate early after ICH may predict for poorer outcomes. A potential mechanism of association between brain and heart was revealed in a work of Wang D, which indicated that autonomic dysfunction characterized by a higher heart rate and blood pressure level after ICH correlated to a poorer functional outcome and increased mortality[18]. However, in time series, the association between resting heart rate (RHR) for all-cause mortality (ACM) in patients with ICH in the intensive care unit (ICU) has not been clearly mentioned or elucidated.

By utilizing data from MIMIC-IV database, we aimed to investigate the association between RHR levels and ACM in this cohort and to fill the current gap in knowledge regarding the predictive effect of RHR for outcomes in patients with ICH to help with health care management and improve clinical outcomes ultimately.

## Materials and methods

### Study population

We conducted a retrospective study involving health-related data from accessing the MIMIC-IV (version 2.2) database which was created and operated by the MIT Computational Physiology Laboratory. The database encompasses a comprehensive and detailed collection of medical records for patients admitted to the intensive care units (ICUs) at Beth Israel Deaconess Medical Center [19]. One of the authors (Xinwei Yao) fulfilled the requirements for database access and was responsible for data extraction (certification number 12928916). This study included patients with ICH according to the International Classification of Diseases, 9th and 10th editions (ICD-9 and ICD-10) from year of 2011-2019. A total of 3,340 adult patients were admitted to the intensive care unit (ICU) at Beth Israel Deaconess Medical Center in Boston. In this cohort, patients diagnosed with ICH were identified on the basis of the following ICD codes: 431, I61, I610, I611, I612, I613, I614, I615, I616, I618 and I619. The exclusion criteria were performed according to certain procedures: (1) participants who were admitted to the ICU more than once; (2) were less than 18 years old; (3) had comprehensive medical data missing; and (4) were diagnosed with severe comorbidities, such as end-stage organ failure or advanced cancer.

Inclusion of a cohort of 1,918 patients was accomplished throughout these criteria. For evaluating the variables related to ICH and for our study purpose, four quartiles were obtained according to average RHR levels, referred to as Q1–Q4.

### Data collection

The data for this study were extracted using Structured Query Language (SQL) (version 13.9.9) in conjunction with Navicat Premium (version 16.1.7) software. The variables chosen for analysis can generally be grouped into five primary categories: (1) demographic variables including age, gender, and race; (2) comorbidities and complications, including hypertension, myocardial infarct, heart failure, intraventricular hemorrhage (IVH), dementia, pulmonary disease, rheumatic disease, mild liver disease, diabetes, paraplegia, and coagulation disorder; (3) vital signs including systolic blood pressure (SBP), diastolic blood pressure (DBP), mean arterial pressure (MAP), pulse oxygen saturation and temperature; (4) scores of clinical rating scales, including the Glasgow Coma Scale (GCS), Sequential Organ Failure Assessment (SOFA), Simplified Acute Physiology Score Ⅱ(SAPS-Ⅱ), Systemic Inflammatory Response Syndrome Score (SIRS) and Oxford acute severity of illness score (OASIS); and (5) laboratory parameters, including hemoglobin, platelets, white blood cell (WBC), bicarbonate, blood urea nitrogen (BUN), calcium, creatinine, chloride, glucose, sodium, potassium, international normalized ratio (INR), partial thromboplastin time (PTT) and prothrombin time (PT).

At the time of admission to the ICU, the follow-up period began, and it ended when death occurred. All laboratory variables and disease severity scores were derived from data documented during the initial assessment following the patient’s admission to the intensive care unit. We examined the data composition of all variables. Variables with data loss greater than 10% were excluded from the study, and those with data loss less than or equal to 10% were supplemented by the multiple imputation of specific variable via SPSS (version 29.0) software.

### Clinical outcomes

The primary endpoints of this observational study were ACM in patients with ICH within follow-up at 30 days, 90 days, and 1 year after ICU admission. Additionally, the survival time was defined as death occurring within a specific period after ICU admission in this study, rather than only identifying whether the patient was excluded at a particular point in the time series.

### Statistical analysis

Subjects were grouped into four quartiles according to their RHR values (Q1–Q4). Continuous variables were tested for normality, and continuous variables with a normal distribution were reported as the mean ± standard deviation (SD); otherwise, they were presented as the median and interquartile range (IQR). Categorical variables were presented as counts and percentages. The t tests or analysis of variance (ANOVA) were utilized for continuous variables with a normal distribution and the Mann–Whitney U tests or Kruskal–Wallis tests were applied for variables with skew distribution. In addition, Pearson’s chi-square test was applied for the comparison of categorical variables across RHR quartiles.

Kaplan-Meier (K-M) survival curves were used to evaluate the incidence of outcomes within groups defined by RHR levels, and the log-rank test was utilized to determine the statistical discrepancies. In addition, covariates were selected through univariate Cox regression analyses with a significance threshold of *P* < 0.05 (detailed in Table Supplementary 1). The confounders were subsequently identified and excluded through mapping and analysis using a directed acyclic graph (DAG), as detailed in Figure Supplementary 1. In addition, the association between RHR and ACM in patients with ICH in the ICU was quantified by multivariate Cox regression models that provided hazard ratios (HRs) and 95% confidence intervals. For model adjustment, well-adjusted models were applied in the multivariate Cox regression analyses: Model 1 (the baseline model, which includes all variables as covariates); Model 2 (which includes age, gender, race, heart failure status, myocardial infarction status, IVH status, dementia status, mild liver disease status, DBP, MAP, SpO_2_, temperature, platelets, bicarbonate, BUN, chloride, and glucose as covariates, selected through univariate Cox regression models); Model 3 (which includes age, gender, race, IVH status, dementia status, mild liver disease status, WBC, platelets, bicarbonate, BUN, chloride, and glucose as covariates, selected through a DAG approach; the procedure is detailed in Figure Supplementary 1).

To thoroughly investigate the correlation between RHR and ACM, restricted cubic splines (RCSs) were utilized to detect potential nonlinear relationships between heightened RHR level and increased risk of ACM with the optimal threshold identified through comprehensive evaluations. Subsequently, a piecewise Cox model was implemented to evaluate the correlation between the risk of ACM and the RHR level across the distinct intervals defined by the identified threshold.

In addition, subgroup analyses and interaction tests were conducted to assess the influence of age (> 60 years or ≤ 60 years), glucose (≥ 200 mg/dL or < 200 mg/dL), gender and the presence of IVH comorbidity on the predictive effect of HRs. If interactions were identified, mediation analysis would be applied for further clarification. The HRs in subgroups were calculated through regression analysis of subgroup data using Model 3.

All analyses were conducted with a significance threshold of *P* < 0.05 (two-tailed) using R and R Studio (version 4.2.2), SPSS (version 29.0) and Graphpad Prism (version 10) software.

### Results Population

The final cohort consisted of 1,918 patients from four racial groups, with a mean age of 67.6 years old (SD=23.6). Among these participants, 970 were male. The participants were divided into four groups based on their RHR levels (bpm): Q1: 40.4-70.2; Q2: 70.2-78.5; Q3: 78.5-88.2; Q4: 88.2-147.9).

### Baseline characteristics

As shown in Table 1, a higher proportion of IVH in Q3 and Q4 (*P* < 0.001) were found, which may suggest that ICH patients with a complication of IVH are more likely to present with higher RHR level. Additionally, a higher proportion of mild liver disease (*P* < 0.001) and diabetes (*P* = 0.004) was observed in Q3 and Q4. The general all-cause mortality rates in the ICU, at 30 days, 90 days, and 1 year were 17.2%, 28.9%, 32.0%, and 37.0% respectively. Furthermore, individuals in Q3 and Q4 were more likely to receive mechanical ventilation (*P* < 0.001) and exhibited with a higher incidence of ACM both in the ICU, at 30 days, 90 days, and 1 year post-ICH (*P* < 0.001). Other baseline characteristics are detailed in Table 1.

**Table 1.**
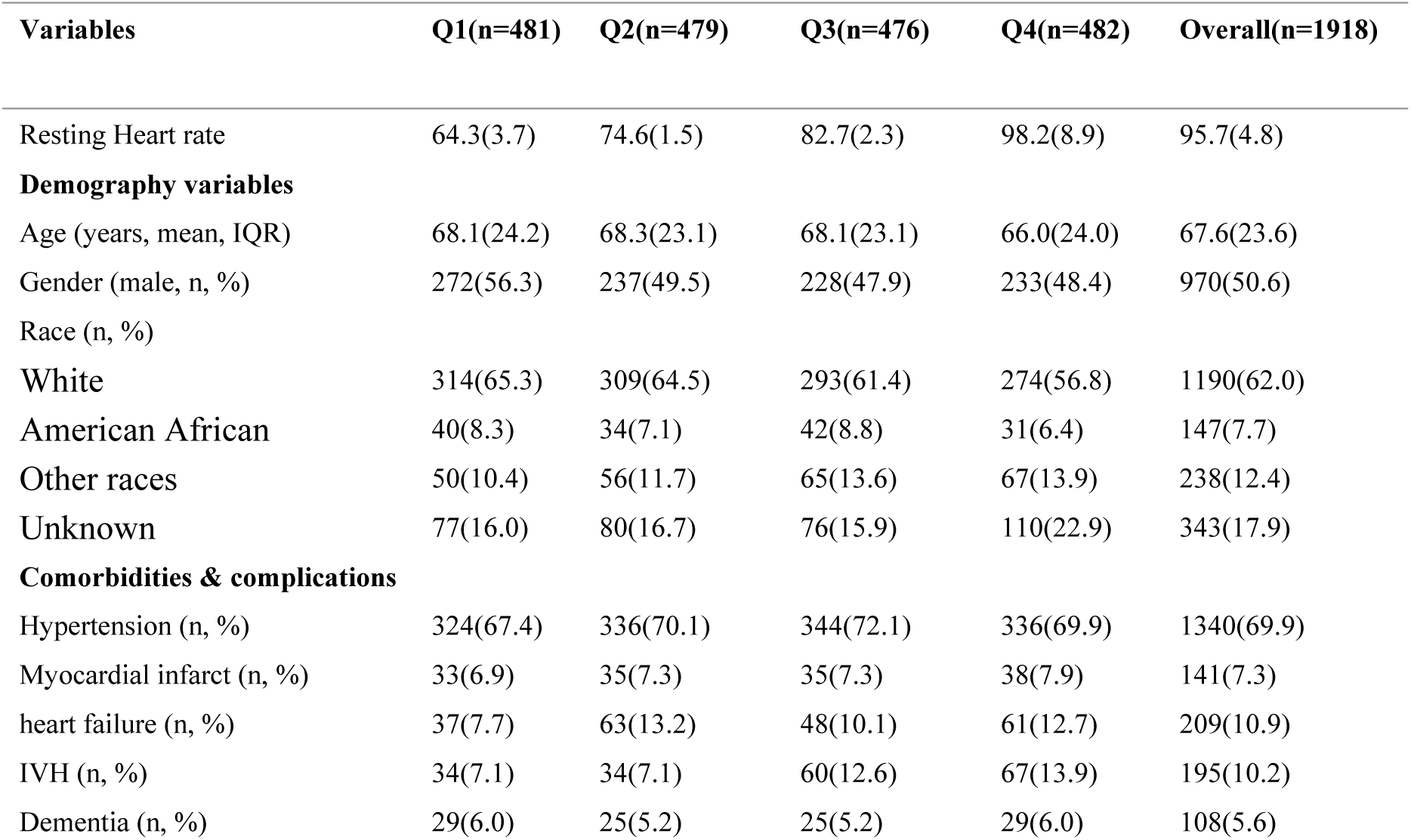
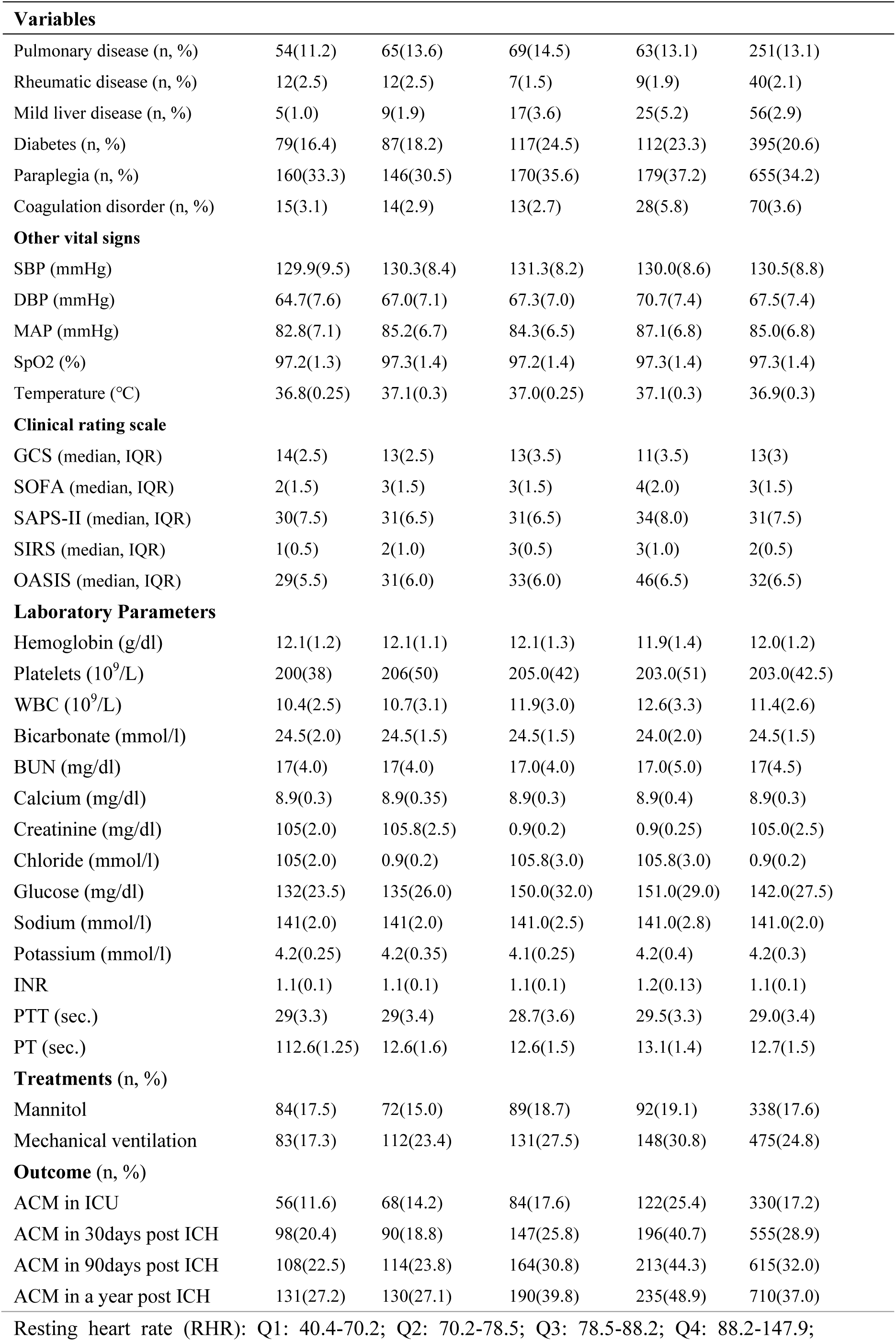
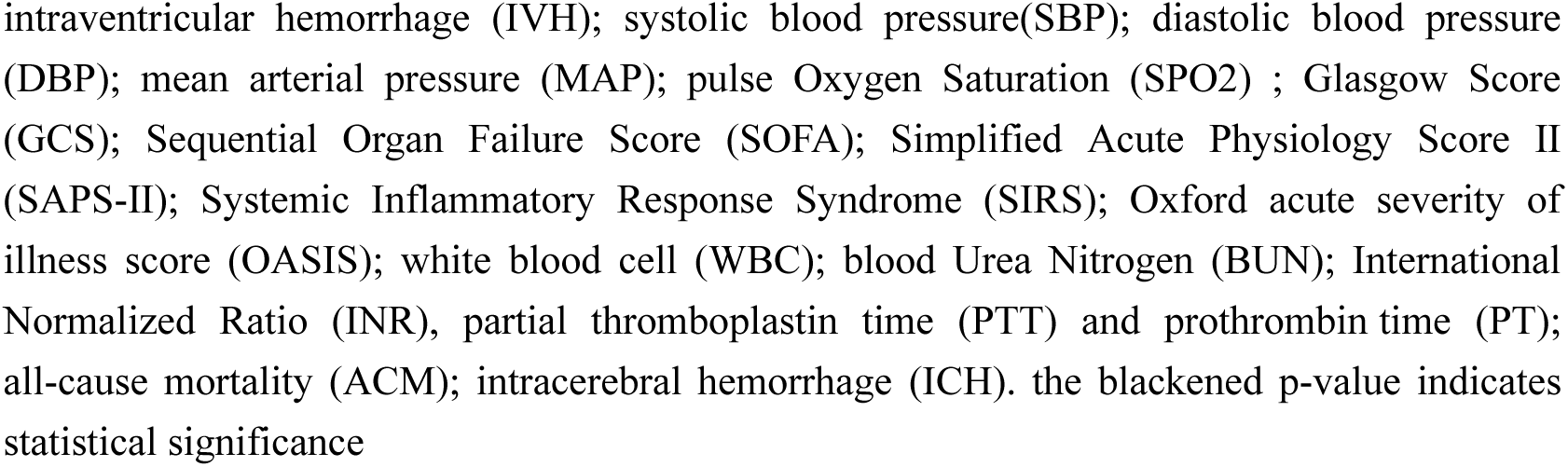
Characteristics and outcomes of ICH participants categorized by resting heart rate.

### Association between RHR and ACM in patients with ICH in the ICU

K-M survival curves were used to evaluate the incidence of ACM across different RHR quartiles over time, as shown in Figure 2 and Table 2. Consistent trends of increasing ACM incidence were observed across all quartiles with longer follow-up durations. Notably, the ACM trend curves of individuals in Q1 and Q2 didn’t show significant difference throughout the time series, while those in Q3 and Q4 both showed significant difference from Q1 and Q2, suggesting that ICH patients with higher RHR may face an elevated risk of ACM over time.

**Figure 1.**
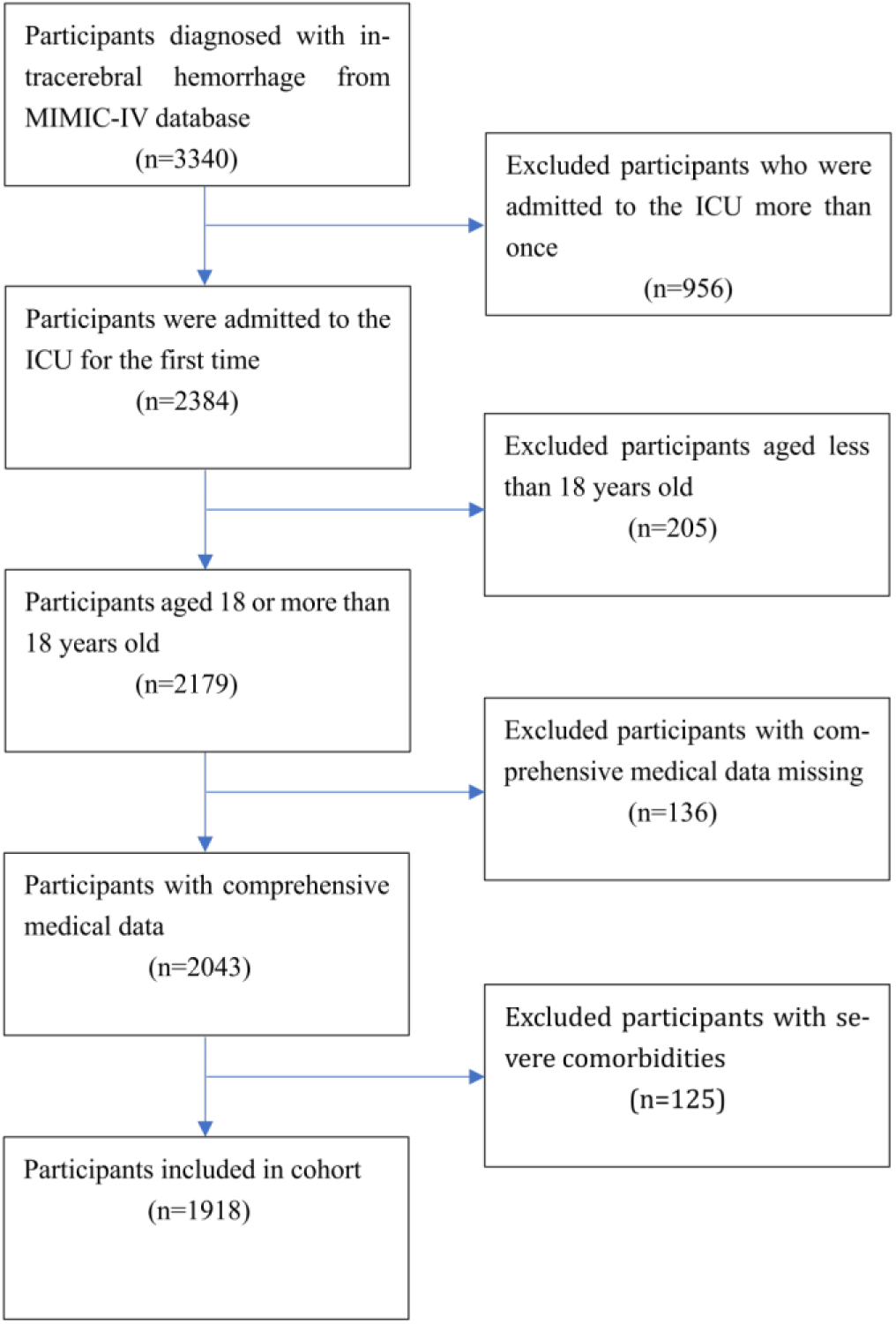
An inclusion and exclusion flowchart; Medical Information Mart for Intensive Care-IV (MIMIC-IV); intensive care unit (ICU).

**Figure 2.**
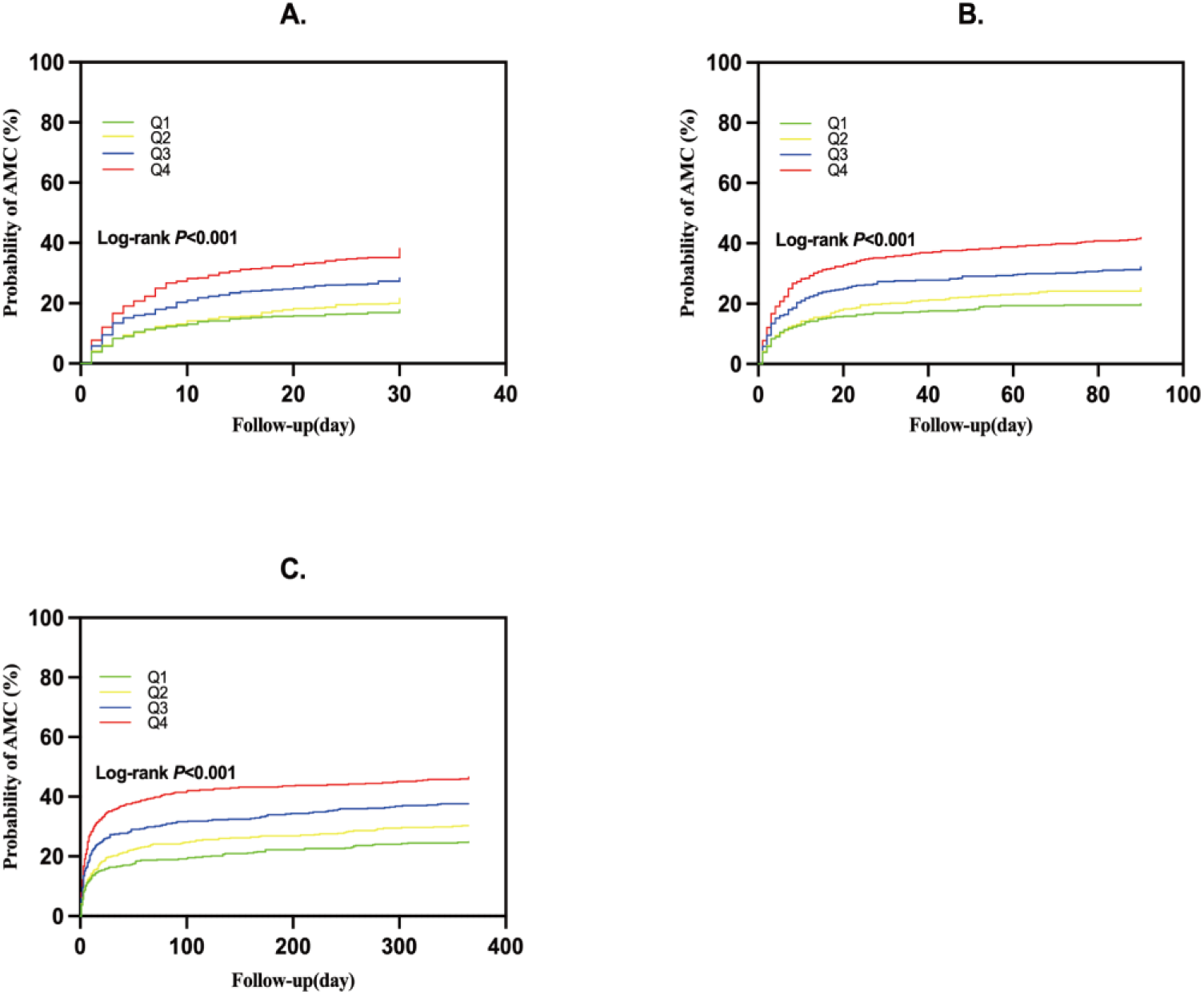
Kaplan-Meier survival curves forall-cause mortality (ACM) in 30 days (A.), 90 days (B.), one year (C.).

**Table 2.** Paired comparisons of the Log-ranks tests of K-M survival analyses.

| Log-rank *P*-value of K-M survival analyses |  |  |  |  |
| --- | --- | --- | --- | --- |
| Quartiles | Q1 | Q2 | Q3 | Q4 |
| ACM in 30 days |  |  |  |  |
| **Q1** | Reference | 0.219 | **< 0.001** | **< 0.001** |
| **Q2** | 0.219 | Reference | **0.017** | **< 0.001** |
| **Q3** | **< 0.001** | **0.017** | Reference | **0.001** |
| **Q4** | **< 0.001** | **<0.001** | **0.001** | Reference |
| ACM in 90 days |  |  |  |  |
| **Q1** | Reference | 0.107 | **< 0.001** | **< 0.001** |
| **Q2** | 0.107 | Reference | **0.014** | **< 0.001** |
| **Q3** | **< 0.001** | **0.014** | Reference | **0.003** |
| **Q4** | **< 0.001** | **< 0.001** | **0.003** | Reference |
| ACM in one year |  |  |  |  |
| **Q1** | Reference | 0.110 | **< 0.001** | **< 0.001** |
| **Q2** | 0.110 | Reference | **0.016** | **< 0.001** |
| **Q3** | **< 0.001** | **0.016** | Reference | **0.001** |
| **Q4** | **< 0.001** | **< 0.001** | **0.001** | Reference |

<sup>2</sup>All-cause mortality (ACM), the blackened *p*-value indicates statistical significance

Since the ACM incidence across all the time points for Q3 was statistically distinct from, and lower than, that of Q4, a concealed nonlinear correlation between rising RHR above the second quartile threshold (>78.5 bpm) and increased risk of ACM emerged. This segmented and significant difference among quartiles prompted the use of restricted cubic splines (RCSs), as detailed in subsequent sections.

After adjustment in multivariable analyses, three multivariate Cox regression models were used to evaluate the association between the RHR level and incidence of ACM in this cohort. As shown in Table 3, the correlation between the RHR level and ACM was not significantly detected in Q1 and Q2. However, when the RHR level valued higher than 78.5 bpm (value of the second quartile of RHR), the correlation between RHR level and ACM was strongly significant (*P* < 0.001), which showed the ACM incidence of Q3-Q4 were approximately 50-100% higher with each 1 quartile increase in RHR compared with that of Q1. In a word, the multivariate Cox regression analyses demonstrated that when the RHR >78.5 bpm, an elevated RHR was independently associated with a higher risk of ACM in whether the short or long time series, which is partly consistent with the findings of the simple K-M survival analyses above.

**Table 3.** Multivariate Cox regression models of ACM in 30 days, 90 days, and one year.

|  | Model 1 |  | Model 2 |  | Model 3 |  |
| --- | --- | --- | --- | --- | --- | --- |
| Quartiles | HR (95% CI) | P-value | HR (95% CI) | P-value | HR (95% CI) | P-value |
| ACM in 30 days |  |  |  |  |  |  |
| Q1 (n=481) | Reference |  | Reference |  | Reference |  |
| Q2 (n=479) | 1.02(0.53-1.96) | 0.961 | 1.17(0.89-1.54) | 0.255 | 1.12(0.86-1.47) | 0.403 |
| Q3 (n=477) | 1.35(0.73-2.50) | 0.333 | 1.51(1.16-1.96) | **0.002** | 1.49(1.15-1.93) | **< 0.001** |
| Q4 (n=481) | 1.70(0.93-3.10) | **0.084** | 2.15(1.64-2.80) | **<0.001** | 2.04(1.59-2.62) | **< 0.001** |
| ACM in 90 days |  |  |  |  |  |  |
| Q1 (n=481) | Reference |  | Reference |  | Reference |  |
| Q2 (n=479) | 1.11(0.84-1.46) | 0.468 | 1.23(0.95-1.59) | 0.123 | 1.16(0.90-1.50) | 0.246 |
| Q3 (n=477) | 1.46(1.12-1.89) | **0.005** | 1.56(1.21-2.01) | **<0.001** | 1.53(1.19-1.96) | **< 0.001** |
| Q4 (n=481) | 2.01(1.56-2.60) | **< 0.001** | 2.22(1.72-2.87) | **<0.001** | 2.07(1.63-2.64) | **< 0.001** |
| ACM in one year |  |  |  |  |  |  |
| Q1 (n=481) | Reference |  | Reference |  | Reference |  |
| Q2 (n=479) | 1.15(0.90-1.45) | 0.262 | 1.20(0.95-1.52) | 0.131 | 1.14(0.91-1.45) | 0.257 |
| Q3 (n=477) | 1.48(1.18-1.86) | < **0.001** | 1.53(1.22-1.93) | <**0.001** | 1.50(1.20-1.88) | < **0.001** |
| Q4 (n=481) | 1.99(1.59-2.50) | < **0.001** | 2.13(1.68-2.69) | <**0.001** | 1.98(1.58-2.47) | < **0.001** |

HR, hazard ratio; CI, confidence interval; Model 1: baseline model including all variables as covariates; Model 2, a model including age, gender, race, heart failure, myocardial infarct, IVH, dementia, mild liver disease, DBP, MAP, SpO<sub>2</sub>, temperature, platelets, WBC, bicarbonate, BUN, chloride, and glucose as covariates through univariate Cox regression analyses; Model 3, a model with exclusion of variables of heart failure, myocardial infarct, DBP, MAP, SpO<sub>2</sub> and temperature. The blackened *p*-value indicates statistical significance

As a segmented correlation between the RHR level and risk of ACM was found in the multivariate Cox regression analyses, restricted cubic splines (RCSs) were used to detect the potential nonlinear correlation. Segmented Cox regression analyses were performed with continuous variable of RHR level as one of the covariates by Model 3. As shown in Figure 3, the RCSs demonstrated that the risk of mortality in the time escalated is in a nonlinear relationship with an elevated RHR in the follow-up at one year (*P*-nonlinearity = 0.036), and when a RHR equal to 90.3 bpm, HRs valued as 1.0 were found in the regression correlation between the RHR level and risk of ACM, which may indicate that a RHR level below 90.3 bpm may be an independent protective factor for ACM in patients with ICH in the ICU, while an elevated RHR level above 90.3 bpm is independently associated with a higher risk of ACM in them.

**Figure 3.**
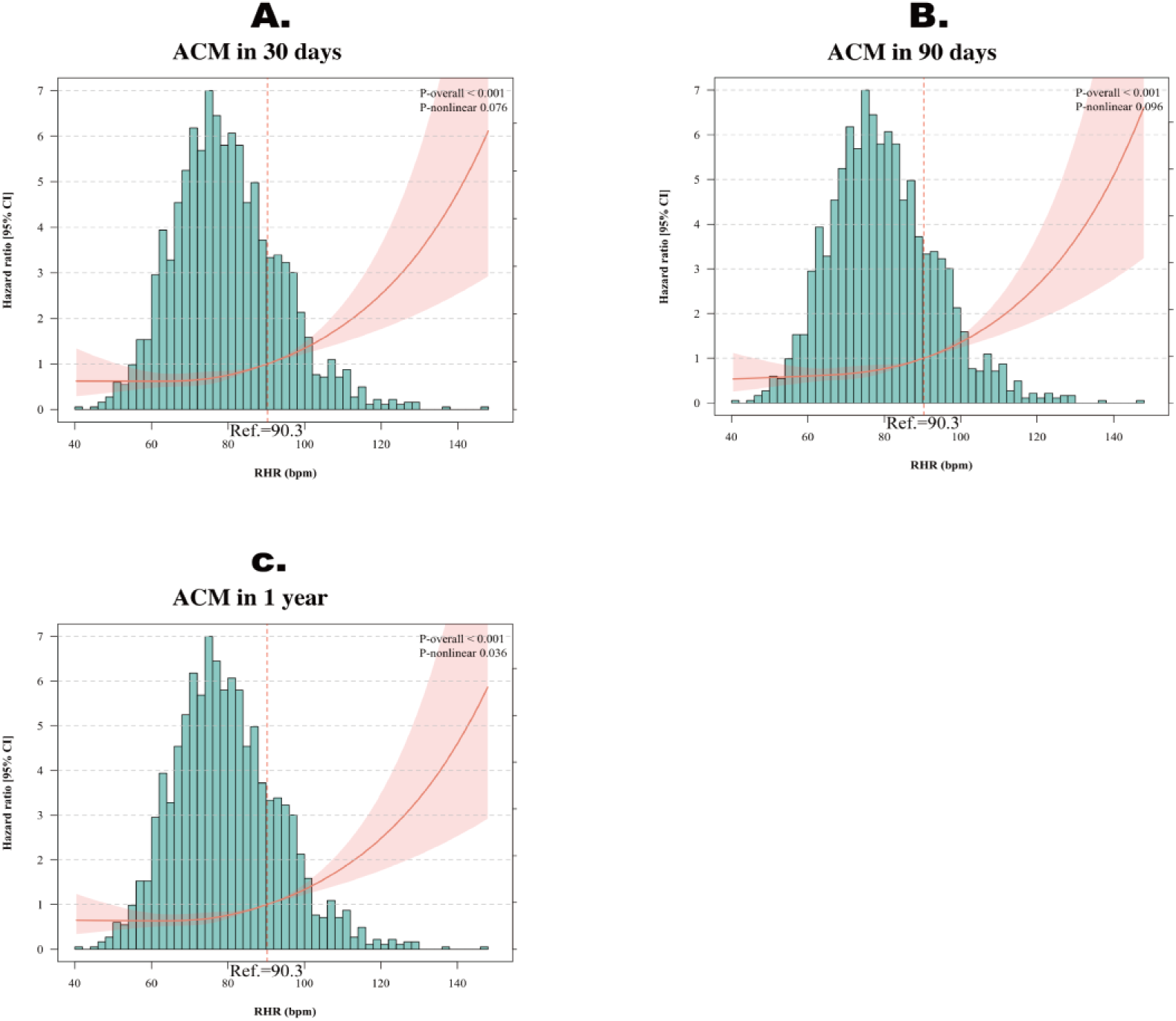
Restricted cubic splines (RCSs) were applied fitting with Cox regression model 3 for 30days (A.), 90days (B.) and one year (C.).

### Subgroup analysis

Subgroups analyses, which included age (>60 years old or ≤60 years old), glucose (≥ 200 mg/dL or < 200 mg/dL), gender and comorbidity of IVH, and interaction tests were applied to assess the potential interaction between these covariates and the effect of the RHR for ACM in patients with ICH in the ICU. As shown in Figure 4, there was an interaction between RHR and age group in these multivariate Cox regression models across all-time series, which showed age of more than 60 years old subgroup had higher HRs between RHR and ACM than those of age less than or equal to 60 years old subgroup. Therefore, to investigate the positive effect of RHR on the incidence of ACM in patients with ICH in the ICU, age was further introduced as a mediator variable into the structural equation model. The bootstrap method provided by Hayes[20] was used to verify the mediating effect of age between the RHR levels and the incidence of ACM. As shown in Figure 5 And Table 4, in this cohort, there was a direct positive correlation between RHR and ACM, while RHR was negatively correlated with ACM through the mediating effect of age, and the negative mediating effects were -0.0036, -0.0041, and -0.0044, with effect ratios of 10.0%, 9.8%, and 12.5% in the time series respectively. In addition, in the subgroup of status of IVH, the regression correlation between the RHR level and ACM was not statistically significant in the follow-up at 30 days. Since IVH is described as one of the severe comorbidities of ICH, it seems understandable that the disease progression of ICH patients combined with IVH could be unpredictable during the early stage of ICH. Moreover, significant correlations between the RHR and ACM were observed in both other two IVH subgroup Cox regression models of 90 days and one year. Generally, the regression models of RHR and ACM in the subgroups were stable and reliable.

**Figure 4.**
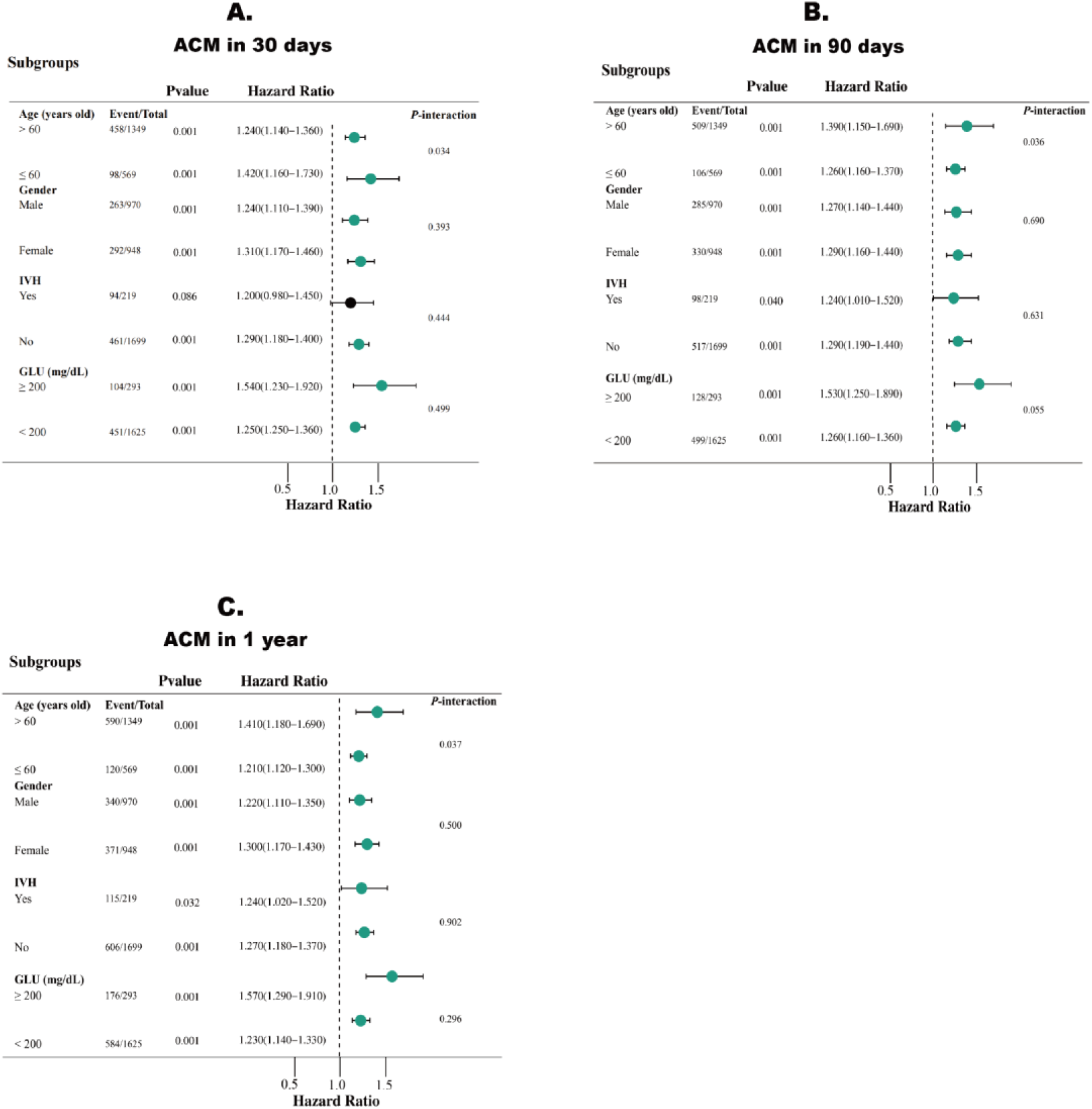
Subgroups analyses and interaction tests in the time series; hazards and confidence intervals were adjusted by Model 3.

**Figure 5.**
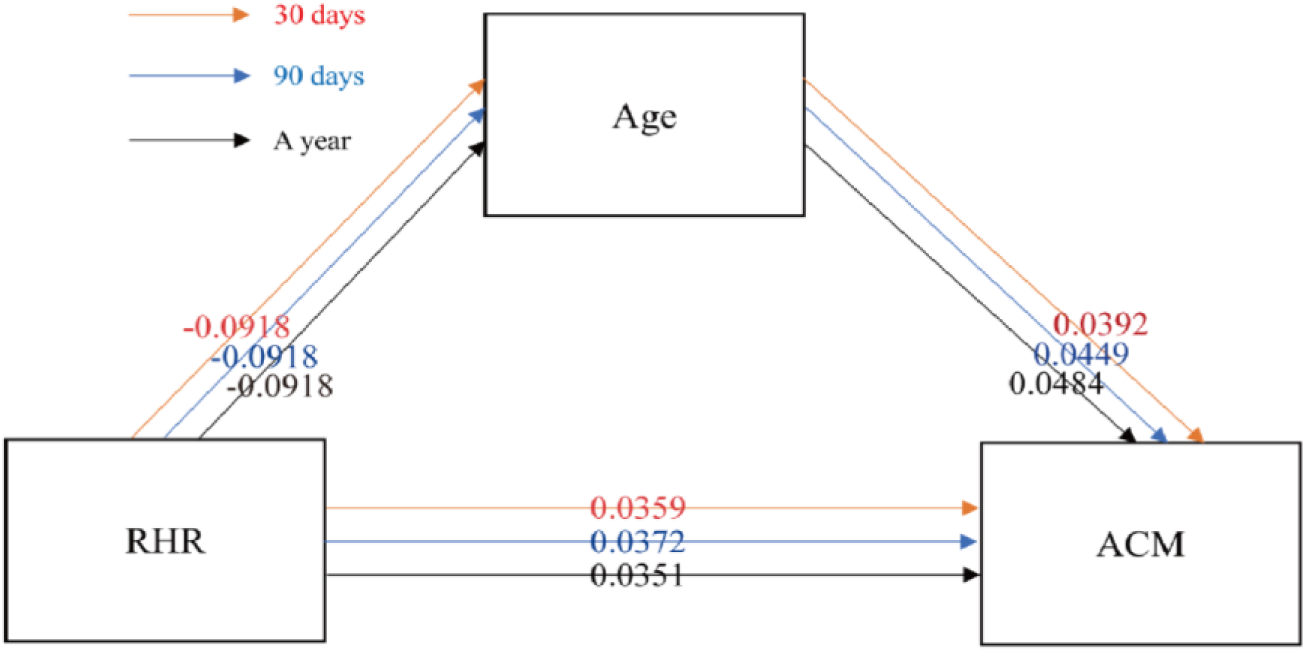
Mediation analyses between resting heart rate (RHR) level and incidence of ACM with age as mediator variable.

**Table 4.**
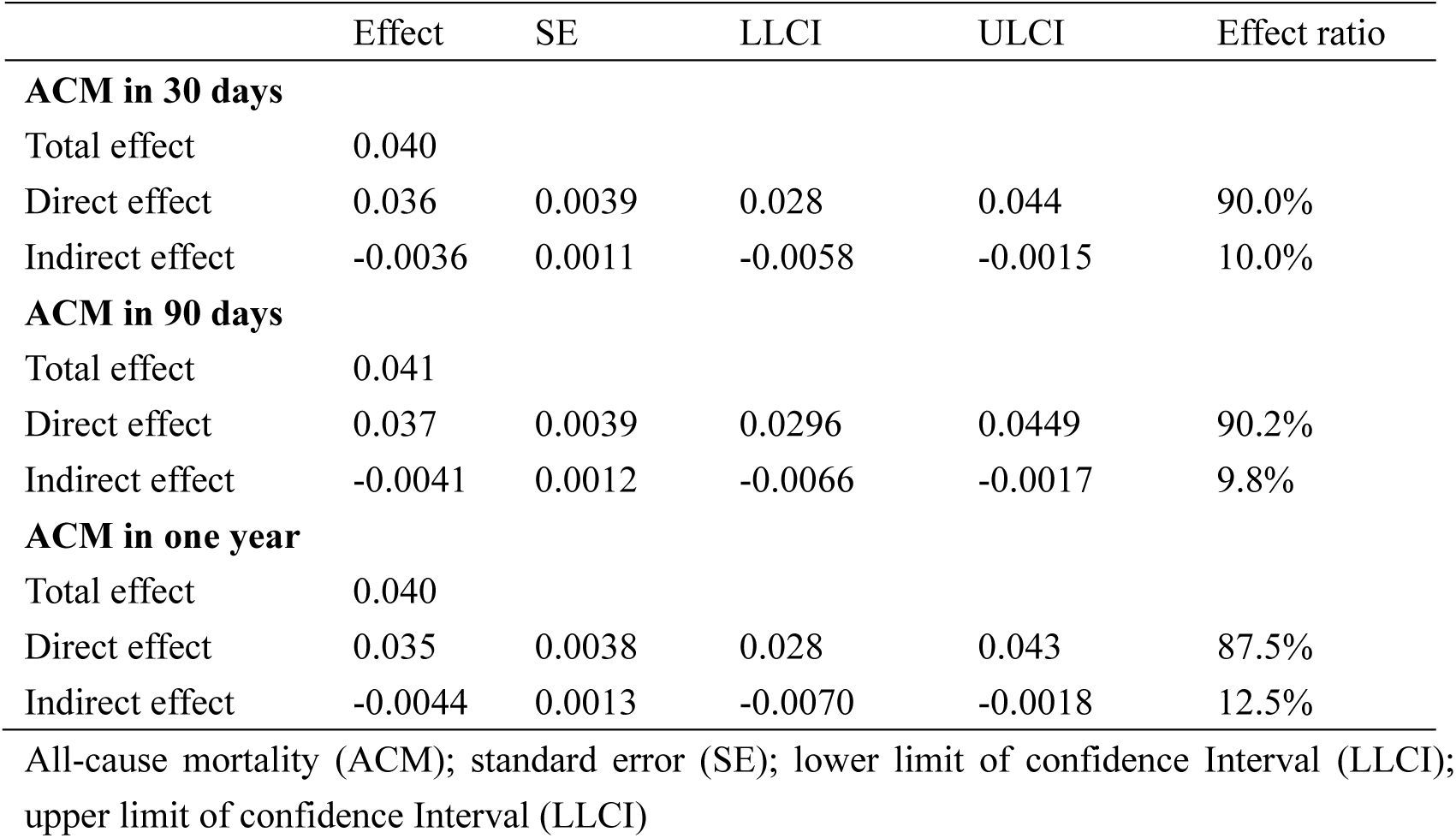
Total effect, direct effect and indirect effect between RHR and ACM.

### Predictive effect of RHR above 90.3 bpm on ACM in patients with ICH in the ICU

As exhibited in Figure 6, as an independent predictive factor for incidence of ACM in patients with ICH in the ICU, RHR above 90.3 bpm showed its reasonably good predictive effect with area under curve (AUC) of more than 0.5.

**Figure 6.**
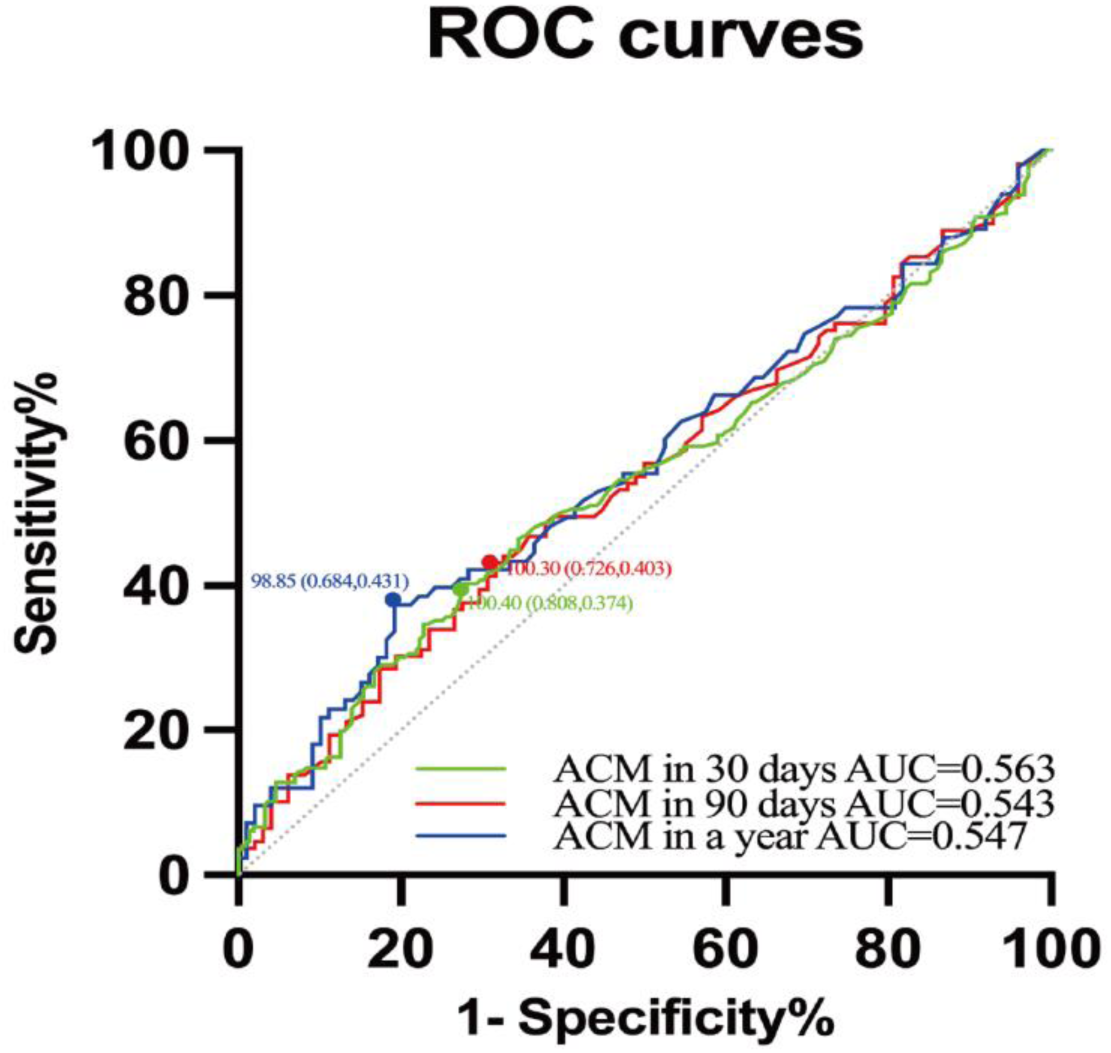
Receiver operating characteristic (ROC) curves; the curves were obtained using RHR above 90.3 bpm as an independent predictor for ACM; highlted spots as cutoff values (specificity, sensitivity) of ROC curves.

## Discussion

There are tight connections between the CNS and cardiovascular system[21]. Heart disease is widely acknowledged as one of the most prevalent causes of stroke[22], meanwhile neurological disorders such as epilepsy, migraine, and cognitive impairment are increasingly linked to coronary artery disease, congenital heart anomalies, valvular disorders, and heart failure[14], [22], [23]. Accumulating evidence indicates that the identification of pathological cardiac status after strokes is important[24–26]. Furthermore, among the various cardiovascular changes following stroke, arrhythmias are notably one of the most detectable yet often overlooked conditions. Additionally, in a retrospective study by Ornella Daniele, arrhythmias were more frequently observed in patients with strokes affecting specific brain regions, suggesting that arrhythmias may be not universally presented poststroke[21], which emphasized that neurologists should be sensitive and cautious to these arrhythmias.

In simple K-M survival analyses (Figure 2, and Table 2), we initially observed a different ACM proportion within the four quartiles. Besides, the marked difference in the ACM proportion between Q3 and Q4 could lead to a further exploration of the nonlinear correlation between a heightened RHR level and increased risk of ACM. Multivariate Cox regression analyses were subsequently conducted with rigorously adjusted models (as shown in Table 3), revealing distinct associations between RHR levels and ACM across quartiles Q1 through Q4. The nonlinear relationship between RHR and ACM was tested via the RCSs (Figure 3). Via the technique of RCSs, a cutoff reference of RHR equal to 90.3 bpm was found, which is considered high yet within a range of normal, and reliable J-shaped curves (*P* < 0.001) were exhibited in all of the time series, which demonstrates the correlation between an elevated RHR level and an increased risk of ACM, and the identification of inflection points in the curves (Figure 3) suggests the possibility of defining an optimal RHR range for patients with ICH in the ICU.

Heart rate, as one of the most important vital signs, has been utilized in clinical score scales and for the prediction of progression and outcome for a long period of time. In a retrospective study [27], 195 patients with ICH who underwent minimally invasive surgery were included and a 12-month follow-up was performed, and the findings of this study indicated that a heart rate of ≥89.50 bpm at discharge was positively associated with mortality at 3 months, whereas a heart rate of ≥92.50 bpm on the third postoperative day was positively associated with mortality at 12 months, which revealed that a heart rate in the normal range could have potential predictive value for the prognosis of certain diseases.

We identified RHR as a promising predictive factor. A new beneficial management strategy involving heart rate control is possible, which can be noninvasive and simple. Then subgroup analyses, as mentioned, revealed interactions between different RHR levels and age groups. The mediation effect analysis (detailed in Figure 5 and Table 4) indicated that RHR level was positively associated with ACM while the RHR level affects ACM through mediating effect of age negatively, and the effect ratios of the indirect effect were significantly lower than those of the direct effect between RHR and ACM, which suggest that the detected interaction between the RHR level and age in this study may be not statistically significant. However, this interaction still needs further elucidation. A large prospective study for exploring the association between RHR and the incidence of atrial fibrillation in a black population, and it demonstrated that an elevated RHR is positively correlated with a higher incidence of atrial fibrillation and ACM, and there was little evidence of effect modification with age[28]. However, another large-scale prospective study with 7976 male participants from Paris reported that individuals with RHR > 90 bpm had a lifespan of 70.27±12.78 years, compared to 79.30±8.43 years in those with RHR < 60 bpm. This study confirmed that the inverse association between resting heart rate and lifespan in human populations, with an additional finding that increases in RHR over time are predictive of higher mortality. Additionally, the results emphasized the importance of monitoring and managing RHR as part of strategies to enhance longevity and public health[29].

In some other studies, the increased blood pressure and heart rate in patients with hemorrhagic stroke was explained by dysfunction of the autonomic nerve system[18], 30–32], so heart rate variability (HRV) was also discussed as a predictive factor for the outcomes of these certain patients. A systematic review revealed that HRV has promising characteristics in prediction of the secondary brain injury, cardiovascular complications and mortality through[33], while in a retrospective cohort study, no variables of HRV were associated with increased 30-day ACM in ICU patients[34]. The predictive value of HRV for disease progression or mortality in the ICU still needs to be evaluated and verified[35].

## Limitations

Although the reliability was shown by adjustment of Cox regression models, in a certain way, we overlooked the integrity of circulation since blood pressure (DBP and MAP included) was excluded as a cofounding factor which may influence both RHR and ACM in a general medical concept. In addition, though the limited value of a RHR >90.3 bpm as an independent predictive factor is shown (as shown in Figure 6.), RHR >90.3 bpm may be considered as a variable in construction of prediction model for ACM of patients with ICH in ICU, and this limitation also underscores the need for high-quality randomized trials and meta-analyses to clarify its role in predicting ACM incidence in patients with ICH. In summary, through this study, the association between the elevated RHR and increased risk of ACM in patients with ICH in the ICU was demonstrated and validated.

## Conclusion

A RHR below 90.3 bpm may serve as an independent protective factor against all-cause mortality in patients with intracerebral hemorrhage in the intensive care unit. In contrast, an elevated RHR exceeding 90.3 bpm may be independently associated with an increased risk of all-cause mortality in this population so that patients with ICH can be grouped by RHR levels, which represents different risks of ACM in patients. Ultimately, an implementable method of accurate heart rate control for patients with specific stratification of RHR levels may help with the development of management strategies and improve clinical outcomes.

## Authors’ contributions

Xinwei Yao, Junyu Yang, Min Liu and Chunlong Zhong designed the study. Xinwei Yao extracted, collected, and analyzed data. Xinwei Yao, Tao Zhang, Ying Pang, Chunyu Zhang and Huang Dan prepared tables and figures. Xinwei Yao, Junyu Yang, Siyi Xu and Tongjie Ji reviewed the results, interpreted data, and wrote the manuscript. All authors have contributed to the manuscript and approved the submission.

## Institutional Review Board Statement

The study adhered to the ethical principles outlined in the Helsinki Declaration. The utilization of the MIMIC-IV database has been approved by the review committees of the Massachusetts Institute of Technology and Beth Israel Deaconess Medical Center, and the requirement for additional ethical approval and informed consent were waived as the data is publicly accessible via the MIMIC-IV database.

## Informed Consent Statement

Not applicable.

## Data Availability Statement

The datasets used and/or analyzed during the current study are available from corresponding author on reasonable request.

## Acknowledgements

We gratefully thank Dr. Jie Liu of Department of Vascular and Endovascular Surgery, Chinese PLA General Hospital & Physician-Scientist Center of China for his contribution to the statistical support, study deign consultations and comments regarding the manuscript.

## Abbreviations

MIMIC-IV: Medical Information Mart for Intensive Care-IV
ICH: Intracerebral hemorrhage
ICU: Intensive care unit
RHR: Resting heart rate
bpm: Beats per minute
ACM: All-cause mortality
ICD: International Classification of Disease
RCS: Restricted cubic splines
GBD 2021: Global burden of disease 2021
CNS: Central nervous system
IVH: Intraventricular hemorrhage
SBP: Systolic blood pressure
DBP: Diastolic blood pressure
MAP: Mean arterial pressure
SpO_2_: Saturation of pulse oxygen
GCS: Glasgow Coma Scale
SOFA: Sequential organ failure assessment
SAPS-II: Simplified acute physiology score Ⅱ
SIRS: Systemic inflammatory response syndrome score
OASIS: Oxford acute severity of illness score
WBC: White blood cell
BUN: Blood Urea Nitrogen
INR: International normalized ratio
PTT: Partial thromboplastin time
PT: Prothrombin time
SD: Standard deviation
IQR: Interquartile range
DAGs: Directed acyclic graphs
HR: Hazards ratio
SE: Standard error
LLCI: Lower limit of confidence Interval
ULCI: Upper limit of confidence Interval
HRV: Heart rate variability

## Supplementary Materials

**Figure S1.**
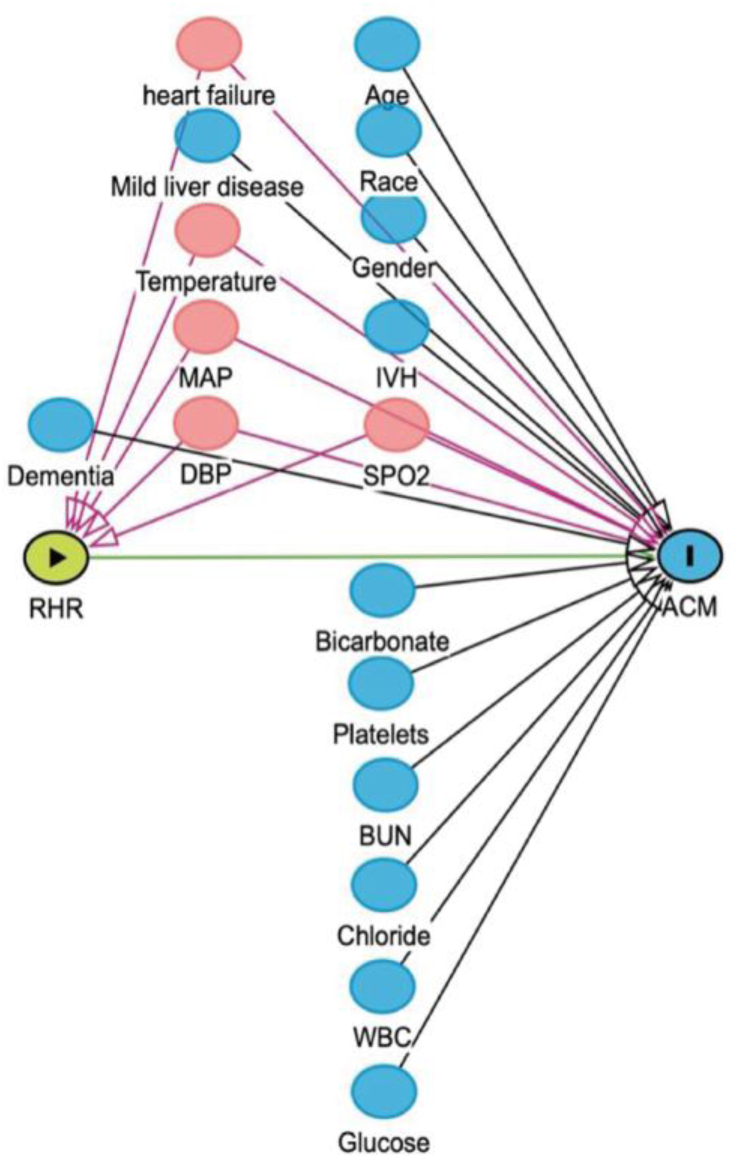
Directed acyclic graphs (DAGs) for selection of cofounding factors; intraventricular hemorrhage (IVH); systolic blood pressure (SBP); diastolic blood pressure (DBP); mean arterial pressure(MAP); pulse Oxygen Saturation (SPO2); white blood cell (WBC); blood urea nitrogen (BUN); red buttons: confounding factors which were excluded from multivariate Cox regression model 2; blue buttons: variables that may influence ACM independently and were included in Cox regression models.

**Table S1.** Univariate Cox regression analyses for covariates selection.

| variables | ACM on 30days | ACM on 90days | ACM in a year |
| --- | --- | --- | --- |
|  | *p*-value (< 0.05) | *p*-value (< 0.05) | *p*-value (< 0.05) |
| **Age** | **<0.001** | **<0.001** | **<0.001** |
| **male** | 0.081 | **0.016** | 0.055 |
| **Race** | **<0.001** | **<0.001** | **<0.001** |
| **Hypertension** | 0.343 | 0.168 | 0.325 |
| **Myocardial infarct** | 0.322 | 0.063 | 0.177 |
| **heart failure** | **0.046** | **0.029** | **0.023** |
| **IVH** | **<0.001** | **<0.001** | **<0.001** |
| **Dementia** | **<0.001** | **<0.001** | **<0.001** |
| **Pulmonary disease** | 0.496 | 0.772 | 0.463 |
| **Rheumatic disease** | 0.201 | 0.296 | 0.595 |
| **Mild liver disease** | **0.027** | **0.023** | **0.007** |
| **Diabetes** | 0.170 | 0.170 | 0.632 |
| **Paraplegia** | 0.988 | 0.957 | 0.795 |
| **Coagulation disorder** | 0.285 | 0.567 | 0.530 |
| **SBP** | 0.190 | 0.247 | 0.308 |
| **DBP** | **<0.001** | **<0.001** | **<0.001** |
| **MAP** | **<0.001** | **<0.001** | **<0.001** |
| **SpO2** | **<0.001** | **<0.001** | **0.038** |
| **Temperature** | **0.003** | **0.007** | **0.018** |
| **Hemoglobin** | 0.558 | 0.677 | 0.748 |
| **Platelets** | 0.204 | 0.087 | **0.048** |
| **WBC** | **0.034** | 0.061 | 0.158 |
| **Bicarbonate** | 0.084 | **0.042** | **0.007** |
| **BUN** | **0.002** | **<0.001** | **0.011** |
| **Calcium** | 0.471 | 0.576 | 0.614 |
| **Creatinine** | 0.910 | 0.881 | 0.889 |
| **Chloride** | **0.047** | 0.062 | 0.110 |
| **Glucose** | **<0.001** | **<0.001** | **<0.001** |
| **Sodium** | 0.856 | 0.639 | 0.905 |
| **Potassium** | 0.349 | 0.325 | 0.599 |
| **INR** | 0.801 | 0.636 | 0.814 |
| **PTT** | 0.690 | 0.458 | 0.222 |
| **PT** | 0.805 | 0.895 | 0.901 |

intraventricular hemorrhage (IVH); systolic blood pressure (SBP); diastolic blood pressure (DBP); mean arterial pressure (MAP); pulse Oxygen Saturation (SPO2); white blood cell (WBC); blood urea nitrogen (BUN); International Normalized Ratio (INR); partial thromboplastin time (PTT) and prothrombin time (PT).the blackened p-value indicates statistical significance.

